# Bayesian sequential approach to monitor COVID-19 variants through positivity rate from wastewater

**DOI:** 10.1101/2023.01.10.23284365

**Authors:** J. Cricelio Montesinos-López, Maria L. Daza–Torres, Yury E. García, César Herrera, C. Winston Bess, Heather N. Bischel, Miriam Nuño

## Abstract

Trends in COVID-19 infection have changed throughout the pandemic due to myriad factors, including changes in transmission driven by social behavior, vaccine development and uptake, mutations in the virus genome, and public health policies. Mass testing was an essential control measure for curtailing the burden of COVID-19 and monitoring the magnitude of the pandemic during its multiple phases. However, as the pandemic progressed, new preventive and surveillance mechanisms emerged. Implementing vaccine programs, wastewater (WW) surveillance, and at-home COVID-19 tests reduced the demand for mass severe acute respiratory syndrome coronavirus 2 (SARS-CoV-2) testing. This paper proposes a sequential Bayesian approach to estimate the COVID-19 positivity rate (PR) using SARS-CoV-2 RNA concentrations measured in WW through an adaptive scheme incorporating changes in virus dynamics. PR estimates are used to compute thresholds for WW data using the CDC thresholds for low, substantial, and high transmission. The effective reproductive number estimates are calculated using PR estimates from the WW data. This approach provides insights into the dynamics of the virus evolution and an analytical framework that combines different data sources to continue monitoring the COVID-19 trends. These results can provide public health guidance to reduce the burden of future outbreaks as new variants continue to emerge. The proposed modeling framework was applied to the City of Davis and the campus of the University of California Davis.

## 1 Introduction

Effectively monitoring the evolution of the COVID-19 pandemic and controlling the spread of disease remains a major public health challenge. The rapid evolution of SARS-CoV-2, the virus that causes COVID-19, and variable community responses to public health interventions involving sustained behavior change, stay-at-home restrictions, face coverings, testing, and vaccination. Statistical and mathematical models are an important component of effective monitoring systems to track COVID-19 cases, hospitalizations, and deaths (Capistran et al., 2021; Giordano et al., 2020). However, classic models in epidemiology are limited. It is necessary to develop monitoring systems that capture the complex dynamics of the evolving pandemic (Daza-Torres et al., 2022; Engbert et al., 2020). Good data availability and quality, which have also changed over time, are essential in mathematical and statistical model development (Jewell et al., 2020; Shinde et al., 2020). Mass testing was an essential control measure for curtailing the burden of COVID-19, particularly during its early phases. As the pandemic progressed, new interventions and monitoring strategies surged, including vaccine programs, wastewater (WW) surveillance (Ahmed et al., 2020; Huisman et al., 2022), and at-home COVID-19 tests (Rader et al., 2022).

Mass testing campaigns, contact tracing, isolation, and mobility restrictions made it possible to estimate the burden of disease during the early phases of the pandemic (Bassi et al., 2021; Yang et al., 2020). However, the return to “normal” was accompanied by a decrease in COVID-19 clinical testing programs and an increase in at-home diagnostic tests. These changes created limits on the utility of individual case data for public health decision-making. Using the number of confirmed cases to determine the prevalence of disease in a community can provide biased information since observed cases depend on the tests conducted, strategies to sample people, and the timing of case detection (Böttcher et al., 2022; Fenton et al., 2020). Therefore, without more specifications, the number of positive cases may not be an appropriate indicator to monitor disease burden. Positivity rate (PR) has been shown to be a better indicator for disease spread than confirmed cases because it considers both tests conducted and cases detected (Dallal et al., 2021; Montesinos-López et al., 2021).

During the COVID-19 pandemic, public health officials commonly used the PR to infer the adequacy of population-level testing and the extent of SARS-CoV-2 transmission in a population (Dallal et al., 2021; Fenga and Gaspari, 2021; Furuse et al., 2021). A low PR indicated low viral prevalence and a testing program with sufficient surveillance capacity. In contrast, a higher PR suggested a low number of tests being conducted, suggesting that many infected individuals were undetected (Dowdy and D’Souza, 2020; Fernandez-Cassi et al., 2021; Mercer and Salit, 2021).

At the beginning of the pandemic, the World Health Organization (WHO) recommended a PR threshold of 5% to declare COVID-19 transmission under control (WHO, 2020). However, as the number of people being tested for COVID-19 changed over time, the PR alone was determined insufficient to assess community-level transmission. Limited levels of testing meant that public health authorities focused on passive case-finding (i.e., only those considered most likely to be infected due to symptoms or contacts were tested). As a result, PR tended to be artificially high, and models using PR as input tended to overestimate the proportion of people infected. This is contrary to models based only on observed cases, which may underestimate the COVID-19 prevalence (Daza-Torres et al., 2023). Despite these limitations, PR can still provide a reasonable estimate of the extent of an outbreak if PR is combined with additional information.

Public health authorities are turning to wastewater-based epidemiology (WBE) as an alternative strategy for less-biased population-level surveillance of SARS-CoV-2 RNA. WBE uses biomarkers in WW to monitor trends in community-level health indices. WBE methods have been used to detect changes in drug consumption (Zuccato et al., 2005), dietary patterns (Choi et al., 2019), and the circulation of pathogens like poliovirus and norovirus (Asghar et al., 2014). SARS-CoV-2 RNA in WW correlates well with reported cases of COVID-19 (Ai et al., 2021; Daza-Torres et al., 2023; Huisman et al., 2022). However, some studies have shown that the relationship between WW and COVID-19 clinical cases varies over time (D’Aoust et al., 2022; Xiao et al., 2022). This relationship is affected by testing practices and the emergence of new variants (Frampton et al., 2021; Graham et al., 2021; Xiao et al., 2022). The work of (Xiao et al., 2022) showed that increments in the ratio of WW signal and reported daily new positive clinical tests could be attributed to insufficient testing in the general population (i.e., insufficient testing to capture the exponential growth of actual COVID-19 cases). These findings suggest that models assuming observed cases as input will tend to underestimate the prevalence of COVID-19, as was noted in (Daza-Torres et al., 2023).

In this paper, we correlate WW with PR to monitor COVID-19 trends and to help overcome the limitations of relying only on clinical cases. We pose an adaptive scheme to model the non-autonomous nature of the prolonged COVID-19 pandemic. The PR is modeled through a sequential Bayesian approach (Daza-Torres et al., 2022) with a Beta regression using the WW viral loads as a covariable. The resulting model allows us to compute a PR based on WW measurements and incorporates changes in viral transmission dynamics through an adaptive scheme. The PR estimate is used to calculate values for WW data that correspond to PR thresholds using criteria proposed by the United States Centers for Disease Control (CDC) in 2021 Christie et al. (2021). The PR thresholds indicated low transmission for a *PR* ≤ 0.05, substantial transmission for a *PR* ∈ (0.08, 0.1), and high transmission for a *PR* ≥ 0.1. Due to uncertainties in relating WW data to COVID-19 case counts, public health authorities typically evaluate trends in WW data to asses changes in infection rates rather than absolute thresholds. However, it may be useful for public health authorities to interpret WW data in terms of PR. Our modeling approach provides insights into the evolution of virus transmission dynamics and a methodology that combines different sources of information to continue monitoring the COVID-19 trends.

The paper is organized as follows: Section 2 describes the data and the modeling approach. Section 4 presents the results 3, and Section 4 discusses our modeling approach’s conclusions and limitations.

## 2 Materials and methods

The analytical framework was developed using data from the City of Davis (Davis) and replicated for the campus of the University of California Davis (UC Davis). The Davis and UC Davis WW collection areas (commonly referred to as sewersheds) are geographically adjacent. The analysis includes laboratory-confirmed incident COVID-19 cases and WW data from July 1, 2021, to July 1, 2022, for Davis and UC Davis. Daily COVID-19 cases and tests for Davis were provided by Healthy Davis Together (HDT) (HDT, 2020). HDT is the community pandemic response program launched in Davis from September 2020 to June 30, 2022, to mitigate the spread of COVID-19. HDT involved a broad set of interventions, including free saliva-based asymptomatic testing with high throughput methods to process large volumes of tests. Testing and cases for UC Davis come from the campus community COVID-19 screening program, which included mandatory completion of bi-weekly asymptomatic tests to access campus facilities. The study site is thus unique compared to most WW surveillance regions in that there was an extraordinarily high number of clinical tests performed in both sewersheds during the study period (Figure 1) for a relatively small population size (425,314 tests were performed over the study period by Davis and 835,785 for UC Davis for approximately 66,799 residents in the combined surveillance regions). The high number of tests resulted in *PR* ≤ 0.05 for the majority of the study period in both locations (Figure 2). These conditions provide a useful context to estimate WW thresholds that correspond to the CDC-defined PR thresholds. The WHO contends that disease dynamics based on case data can be confidently tracked when *PR* ≤ 0.05 (WHO, 2020). Otherwise, when *PR*≥0.05, WW measurements offer a more robust measure of true disease dynamics (Fernandez-Cassi et al., 2021)

**Figure 1:**
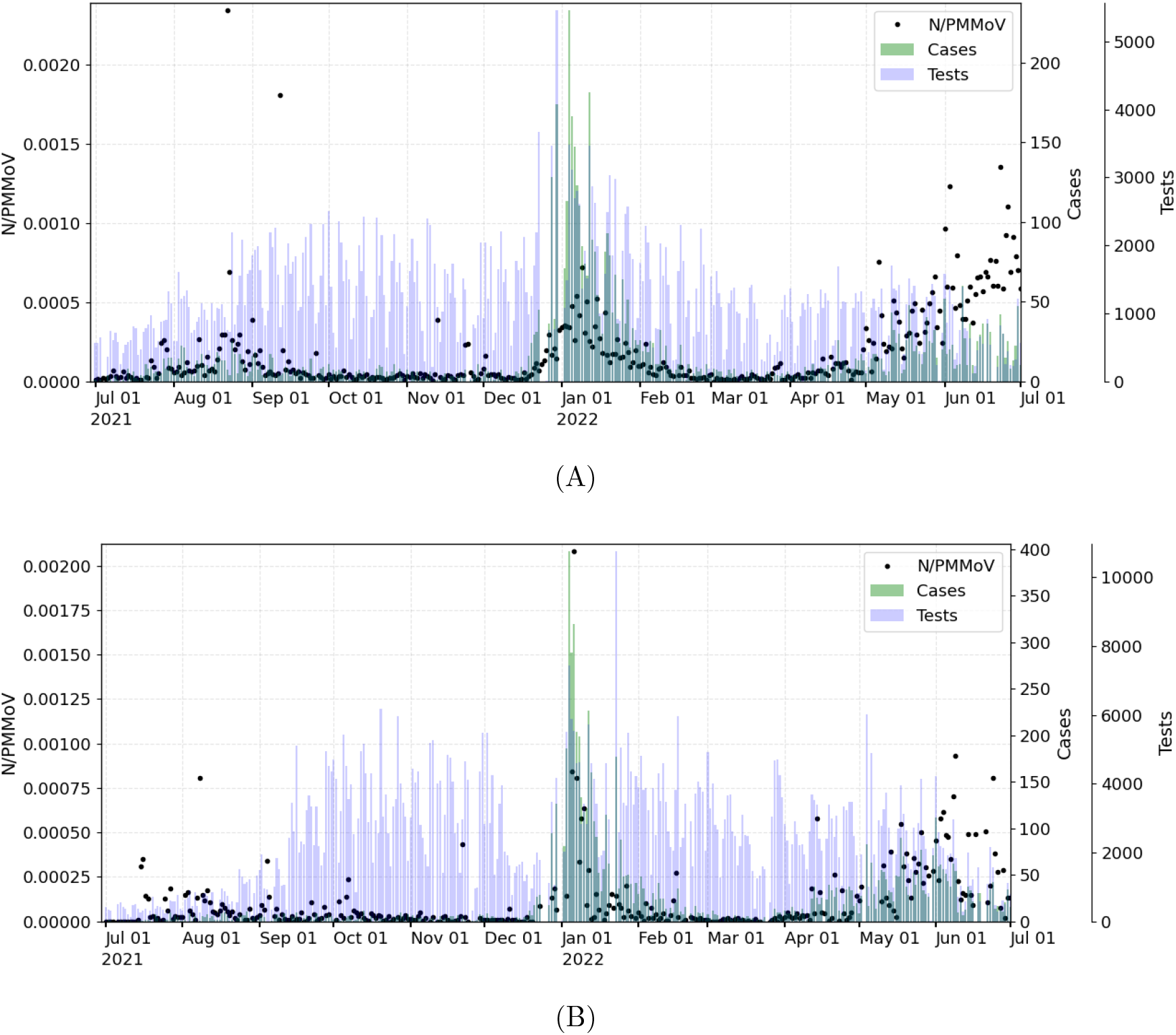
Normalized wastewater data (N/PMMoV), number of COVID-19 tests conducted (Tests), and positive cases (Cases) for (A) Davis and (B) UC Davis.

**Figure 2:**
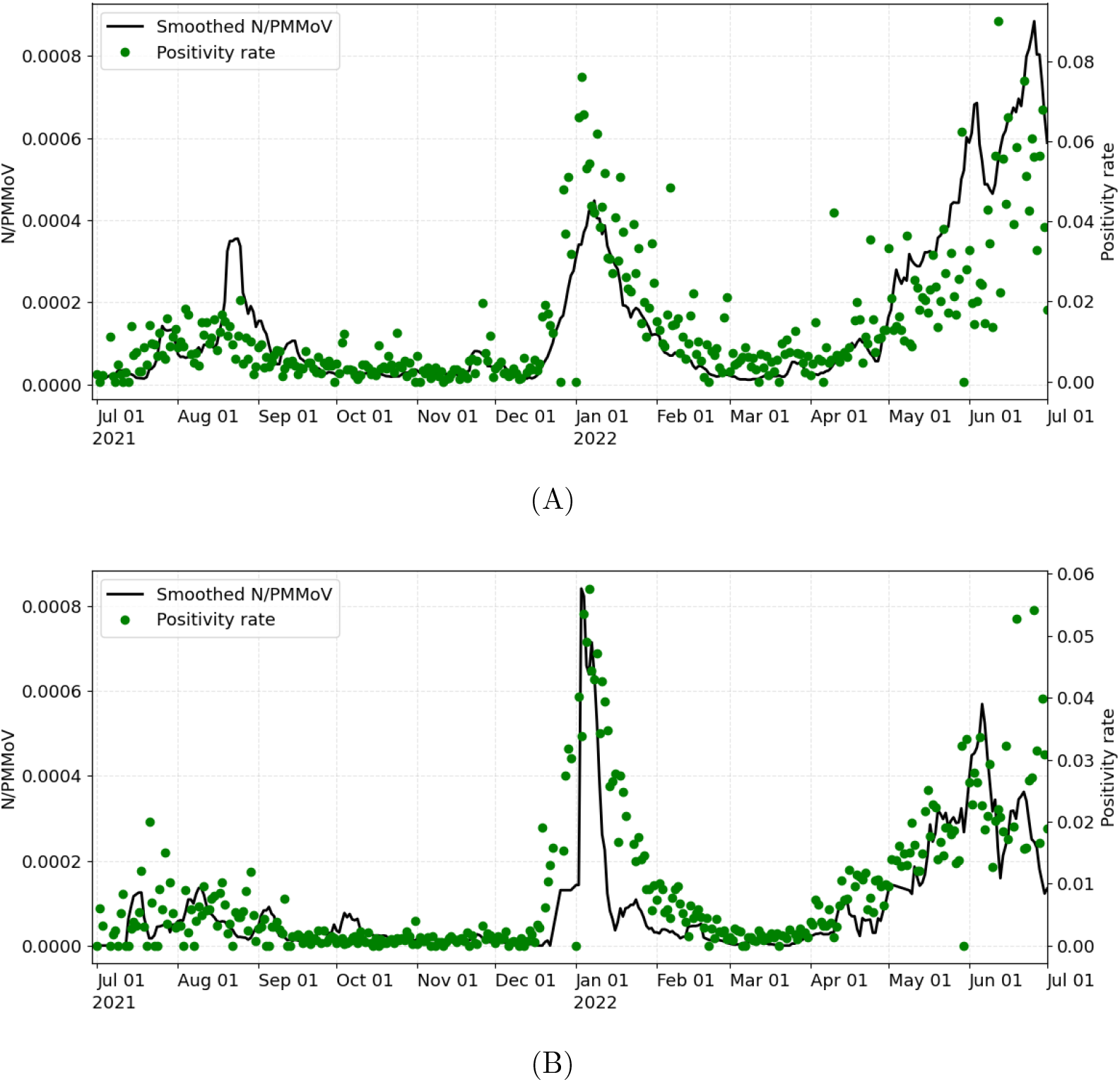
7-day trimmed mean of WW data (Smoothed N/PMMoV) and daily PR for (A) Davis and (B) UC Davis (July 1, 2021 – July 01, 2022).

### 2.1 Wastewater settled solids methods

Wastewater settled solids for the Davis wastewater treatment plant (WWTP) were collected daily from the primary clarifier, transported on the same day of collection to the analytical laboratory, and processed within 24 h as previously described (Wolfe et al., 2021). Wastewater settled solids were obtained from daily composite influent samples from the UC Davis WWTP. Composite influen samples were collected using a refrigerated autosampler (Hach Sigma 900 MAX) located at the WWTP headworks and programmed to collect flow-weighted influent sample volumes every 20 minutes for a total volume of 19 L in 24 hours. Composite influent samples were then transferred to one or two 4L low-density polyethylene containers (LDPE Cubitainers^™^, Thermo Scientific^™^ I-Chem^™^) and stored at 4C prior to settling (up to 6 days of storage). Each 4L sample was pasteurized in a 60C water bath for 45 min immediately prior to settling. Pasteurized influent samples were inverted to mix, poured into a 3-gallon high-density polyethylene conical vessel equipped with a sampling port (FF3G, FastFerment^™^), and left to settle for 2 hours. Settled solids were obtained from either 4L or 8L of influent from a single day (two 4L samples were combined into one settling vessel when 8L were used). Settled solids were collected by dispensing from the bottom of the settling vessel into one or two 50mL polypropylene centrifuge tubes (for 4L or 8L initial volume, respectively). If two tubes of settled solids were obtained, the supernatants were carefully decanted from each, and remaining settled solids were combined. Between sampling episodes, the settling tanks were emptied and the tank and sampling port valves were bleached (10% commercial bleach for 1 hr), rinsed with deionized water, and left to air dry. Samples of settled solids were stored at 4C and subsequently transported on ice in a cooler by a courier to the laboratory.

Samples processing was completed within 8 days of initial sample collection. Sample RNA extraction, purification, and droplet digital reverse transcriptase PCR (ddRT-PCR) followed the same protocol as for the Davis samples. These protocols are described in detail elsewhere (Topol et al., 2021, 2022).

We normalize the SARS-CoV-2 RNA concentration determined (N gene copies per gram dry weight solids) by the concentration of mild pepper mottle virus (PMMoV gene copies per gram dry weight solids) to yield the dimensionless metric, N/PMMoV. The N gene is present in all variants of the virus. PMMoV is a highly abundant RNA virus detected broadly in WW (Rothman et al., 2021; Symonds et al., 2018). PMMoV serves as a process control such that normalization of N by PMMoV for each sample helps correct SARS-CoV-2 concentrations for virus extraction efficiency. PMMoV is also often used to account for variations population size, rainfall, and water usage between different WW collection areas. We expect these latter factors to have less of an effect on N gene concentrations determined herein because water is removed from the WW settled solids samples prior to sample analysis and concentrations are reported in terms of the dry weight of the dewatered solids. Figure 1 shows the normalized N gene concentration.

#### 2.1.1 Smoothed wastewater signal

Given that WW signals are often noise-corrupted, we applied a 7-day trimmed mean for daily WW data to reduce uncertainty and minimize daily fluctuations. The smoothed data is later correlated to raw PR (Figure 2).

### 2.2 Statistical model

We model PR as a Beta distribution using WW viral loads as a covariate, assuming a Bayesian approach. We assume *Y*_*i*_ as the PR at day *i*, defined as the ratio of the number of new positive cases among the number of tests performed at day *i*. Beta regression is a good choice of model for continuous data with response variables expressed as proportions.

The Beta distribution is reparametrized as *Y*_*i*_ *∼ ℬ* (*µ*_*i*_, *ϕ*), with mean *µ*_*i*_ and variance 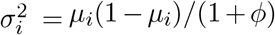, *ϕ* is known as the precision parameter since, for fixed *µ*_*i*_, the larger *ϕ* the smaller the variance of *Y*_*i*_; *ϕ* ^*−*1^ is a dispersion parameter (Ferrari and Cribari-Neto (2004)). The mean *µ*_*i*_ can be expressed as a function of the linear predictor *η*_*i*_ = *β*^*T*^ **x**_*i*_, where *β* is a (*p* + 1)-dimensional vector of unknown regression coefficients (including the intercept), and **x**_*i*_ is the vector of covariates plus a one for the intercept. In this study, only the 7-day trimmed mean of the wastewater data (Smoothed N/PMMoV), denoted as *C*_*i*_, is included in the linear predictor. Thus, the linear predictor is given by *η*_*i*_ = *β*_0_ + *β*_1_*C*_*i*_, and the logit link, the inverse of the logistic function, is used in the Beta regression (i.e., 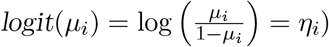.

The aim of this inference problem is to estimate *θ* = (*β*_0_, *β*_1_) from measurements of WW data, **C** = (*C*_1_, *C*_2_, …, *C*_*n*_), and COVID-19 PR, **Y** = (*y*_1_, *y*_2_, …, *y*_*n*_). Thus, the likelihood function for the previous model is given by:

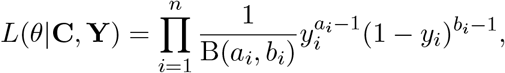

where *a*_*i*_ = *µ*_*i*_*ϕ, b*_*i*_ = *ϕ − a*_*i*_, *µ*_*i*_ = 1*/*(1 + exp (*−η*_*i*_) is the mean (logistic function), *η*_*i*_ = *β*_0_ + *β*_1_*C*_*i*_ is the linear predictor, and *ϕ* is the dispersion parameter.

#### 2.2.1 Bayesian statistical approach

We adopt a Bayesian statistical approach, which is well suited to model multiple sources of uncertainty and allows for incorporating background knowledge of the model’s parameters. In this framework, a prior distribution, *π*_Θ_(*θ*), is required to account for unknown parameter *θ* in order to obtain the posterior distribution. Having specified the likelihood and the prior, we use Bayes’ rule to calculate the posterior distribution,

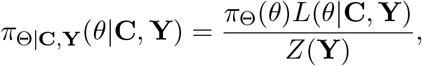

where *Z*(**Y**) = ∫*π*_Θ_(*θ*)*L*(*θ*| **C, Y**)*dθ* is the normalization constant. The posterior distribution is simulated using an existing Markov chain Monte Carlo (MCMC) method, the t-walk algorithm (Christen et al., 2010).

#### 2.2.2 Bayesian sequential method

We adapt the sequential approach proposed in Daza-Torres et al. (2022) to our model to update forecasts over time. The aim is to train the model using only a subset of the most recent data. The forecast is then updated sequentially in a sliding window of data.

We let *L* be the length in days of the period used to train the model. The data window is then moved forward every *n* day as new data becomes available. We set *t*_0_ as the first initial time to start the analysis and the subsequent initial times as *t*_*k*+1_ = *t*_*k*_ + *n*. The training period is taken as [*t*_*k*_, *t*_*k*_ + *L*] and the forecasting period as [*t*_*k*_ + *L, t*_*k*_ + *L* + *F*], see Figure 3.

**Figure 3:**
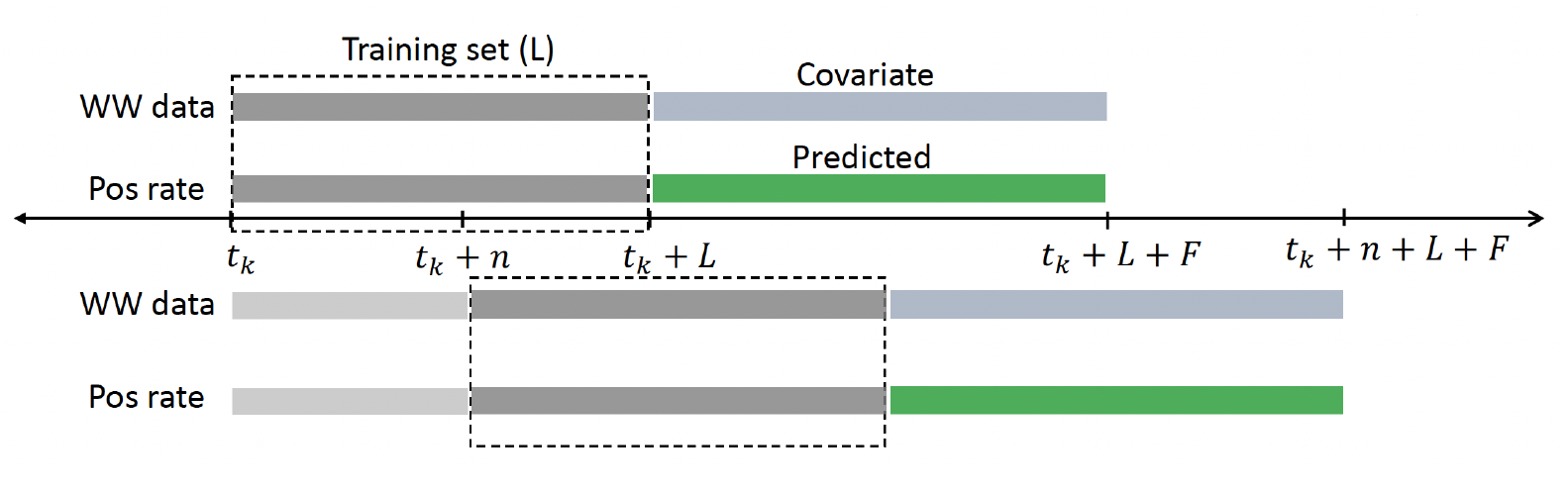
The model is fitted with data from the training period (grey). Then, the estimated parameters are used to predict the forecasting period (green). The training window is then moved *n* days forward. When new data becomes available, we update all forecasts; the latest posterior becomes the newest prior to the next training period.

We denote 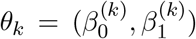 the model parameters to be inferred and the vectors of data as **C**_*k,n*_ = (*C*_*k*_, …, *C*_*k*+*n*_), **Y**_*k,n*_ = (*y*_*k*_, …, *y*_*k*+*n*_) at period *k*. Note that, from the beginning, *θ*_*k*_ is assumed to change in time within each forecast window. If *k* = 0, we postulate a prior distribution 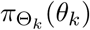 and a likelihood *L*(*θ*_*k*_|**C**_*k,n*_, *Y*_*k,n*_) previously described in Section 2.2. The probabilistic prediction of *y*_*t*_, in the forecasting period *t* ∈ [*t*_*k*_ + *L, t*_*k*_ + *L* + *F*], is obtained by using the estimated parameters through the MCMC method and the WW data concentration (**C**_*k*+*L,F*_).

Afterward, the forecasting window is updated by setting *t*_*k*+1_ = *t*_*k*_ + *n*, with *n* the number of days until the next forecast. In the new training window [*t*_*k*+1_, *t*_*k*+1_ + *L*], we propose a new prior distribution 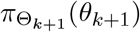 for the model parameters *θ*_*k*+1_ using samples from the posterior distribution obtained in the previous forecast. Finally, we set *k* = *k* + 1 and repeat the process described above to create a new forecast, see Daza-Torres et al. (2022) for implementation details.

Regarding the elicitation of the parameters’ prior distribution for the first forecast, at *k* = 0, we assume a Normal distribution for the parameters 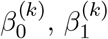, with mean and standard deviation (0, 0) and (1000, 1000), respectively (i.e. noninformative priors). We set *L* to twice the length from symptoms onset to mild disease clinical outcome, namely 30 days, and *F* is chosen to be 10 days. The forecasting was updated every 10 days.

### 2.3 Action thresholds for WW concentration

We let *Y* be the PR, and *C* corresponds to the SARS-CoV-2 RNA concentrations measured in wastewater. Then, the cumulative distribution function of *Y* given *C* is defined as *FY* |*C* (*y*|*c*) := *P* (*Y ≤ y*|*C* = *c*), which represents the probability that PR is less than or equal to *y* given that the SARS-CoV-2 RNA concentration in wastewater is *c*. Henceforth for simplicity, we will use *F* (*y*|*c*) instead of *FY* _|_*C* (*y*|*c*) without loss of generality. The quantile function *F*^*−*1^ of *Y* given *C* = *c* is defined^1^ by

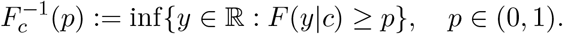

The *p*-quantile of a data set is defined as the value where a *p* fraction of the data is below that value and (1 *− p*) fraction of the data is above that value (e.g., the 0.5 quantile is the median).

Using the CDC thresholds for PR values corresponding to low (*Y* ≤ 0.05), moderate (*Y* ∈ (0.05, 0.08)), substantial (*Y* ∈ (0.08, 0.1)), and high (*Y* ≥ 0.1) transmission and the parameter estimates from the assumed beta regression model, we propose a methodology for estimating WW concentrations associated with PR thresholds at a given point in time. We find the value of WW concentrations, *c*, such that with probability 1 *−α*, the PR is less than the CDC threshold *y∈ {*0.5, 0.8, 0.1*}* (that is, to find *c* such that 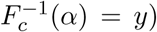. Note that *α* is the precision we set to estimate the threshold. With *α* = 0.05 we are being conservative and choose lower bounds of WW concentration values associated with PR thresholds.

The proposed method for finding these thresholds, assuming that we have simulations of the posterior distribution for the parameter *θ* = (*β*_0_, *β*_1_), is given in Algorithm 1. First, we suggest a search grid for the WW viral load concentration. Then, for each concentration, we simulate the predicted posterior distribution of the PR. Lastly, we calculate the *α*-quantile for each concentration level and find the concentration level that yields the quantile closest to the value of the desired PR threshold.

#### Algorithm 1: Finding threhsold for WW using PR

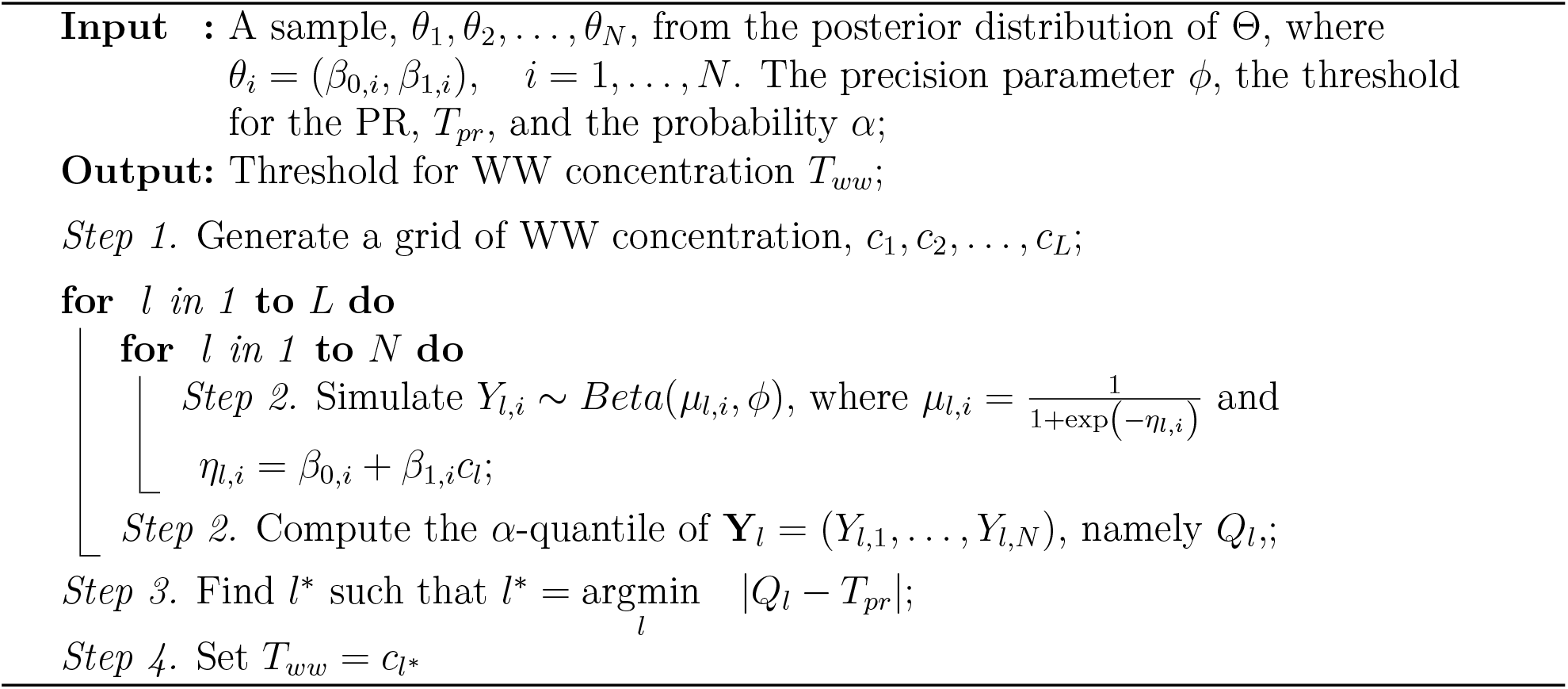

### 2.4 Effective reproductive number

The number of people in a population who are susceptible to infection by an infected individual at any particular time is denoted by *R*_*e*_, the effective reproductive number. This dimensionless quantity is sensitive to time-dependent variation due to reductions in susceptible individuals, changes in population immunity, and other factors. *R*_*e*_ can be estimated by the ratio of the number of new infections (*I*_*t*_) generated at time *t*, to the total infectious individuals at time *t*, given by 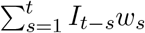, the sum of new infections up to time step *t−* 1, weighted by the infectivity function *w*_*s*_. We implement Cori et al. (2013)’s approach to calculate the *R*_*e*_ from the PR estimated with the WW data. Note that the PR is an estimation of the proportion of infected persons. Therefore, if we multiply the PR by a T value, representing the total number of tests carried out in the study period, we will have an estimate of the incidence, which we can use to compute the *R*_*e*_. Since *R*_*e*_ is a scale-free metric, we should get similar results for different *T* values. We use *T* as the average of the tests carried out in Davis or UC Davis.

## 3 Results

The analytical framework was developed using the City of Davis data and replicated for the University of California, Davis campus.

To display changes in the relation between PR and signal WW across variants, we estimate the PR using the sequential model proposed in Section 2.2.2 and compute the posterior distribution for the parameter *β*_1_ over time (Figures 4-5 top-panel). The results show how the PR and WW signal relation changed, mainly when a new variant emerged. These changes remain similar during the period where the variant is dominant.

**Figure 4:**
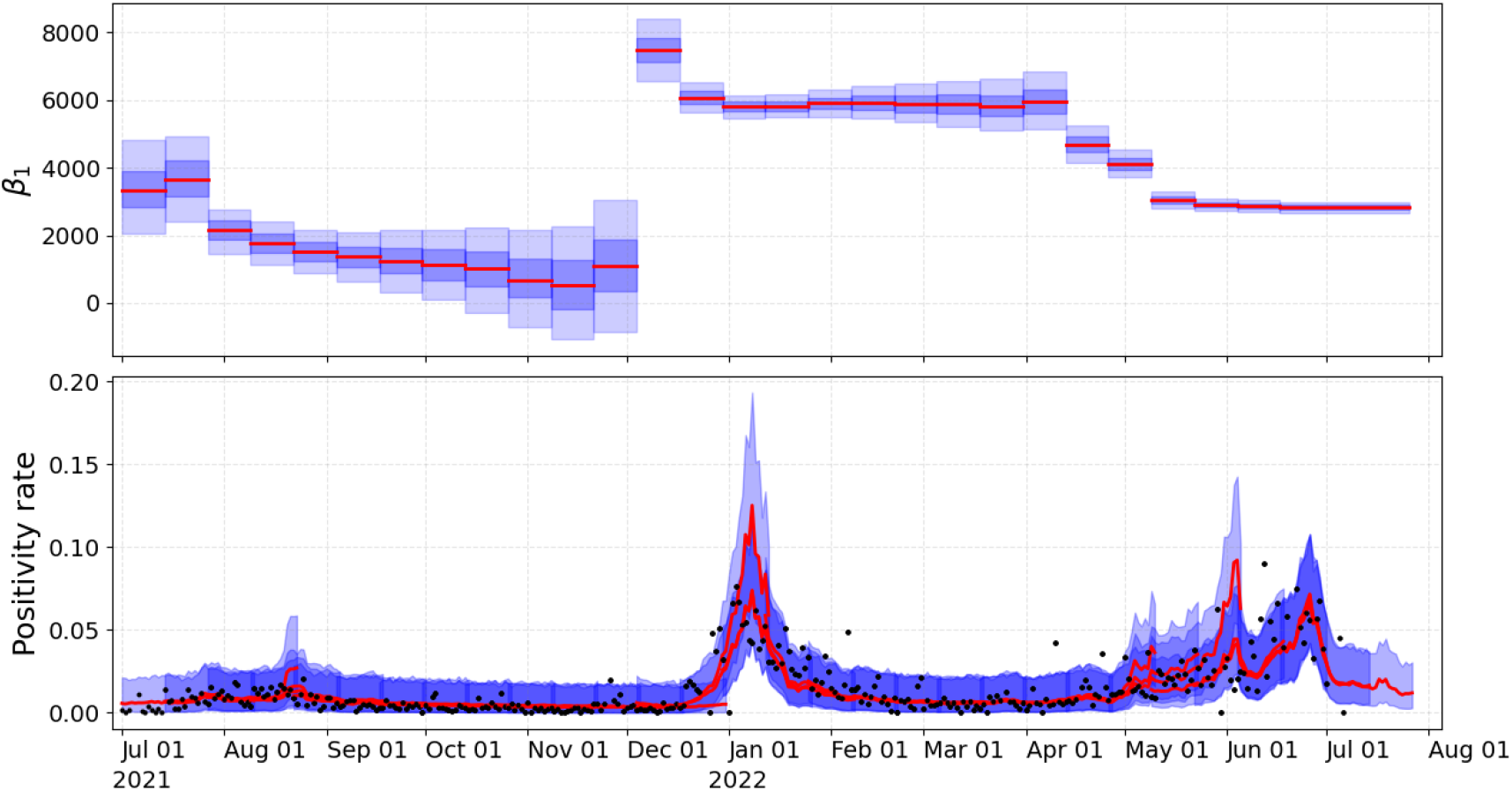
Estimated PR for the City of Davis (lower-panel) and posterior distribution of *β*_1_ (upper-panel). Red-solid line and blue-shadow area describe the mean and SD, respectively.

**Figure 5:**
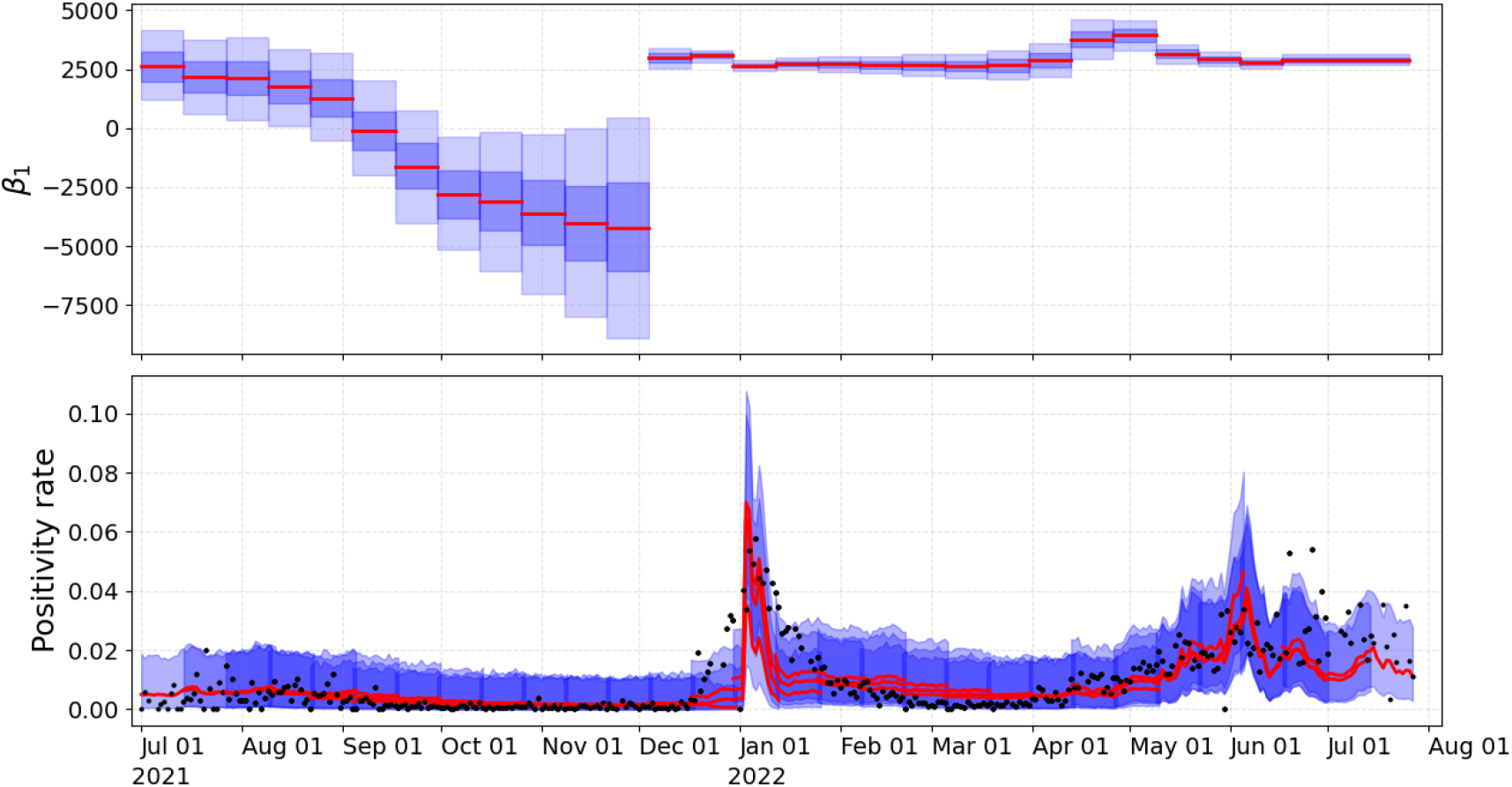
Estimated PR for UC Davis (lower-panel) and posterior distribution of *β*_1_ (upper-panel). Red-solid line and blue-shadow area describe the mean and SD, respectively.

Using the algorithm to compute the thresholds for WW data with the estimated PR in each time window. We did not calculate thresholds for the period of September-December 2021 since the corresponding posterior distribution for *β*_1_ contained zero. In other words, the probability that the *β*_1_ parameter will be zero was positive, suggesting that there is no significant association between WW data and PR (Figures 4-5). We illustrate the estimated thresholds for WW corresponding to low, moderate, substantial, and high transmission thresholds proposed by the CDC (Figure 6). High variability in the threshold estimation coincides with the emergence of new variants. This variability is reduced in the period where the variant is dominant.

**Figure 6:**
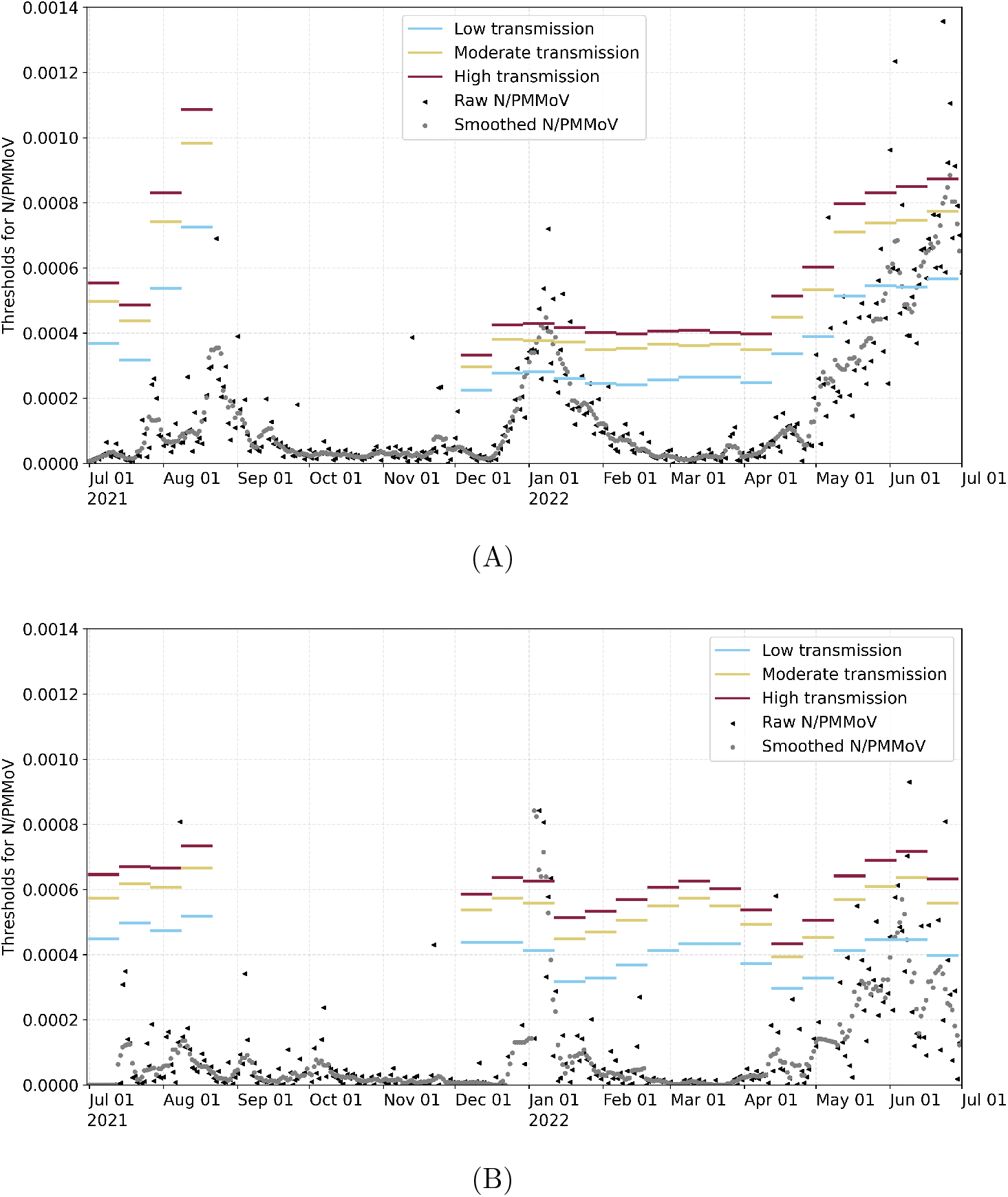
Wastewater thresholds over time for (A) the City of Davis and (B) UC Davis. Blue, yellow, and red horizontal lines correspond to low, moderate, and high transmission thresholds, respectively. Raw (black triangles) and Smoothed (grey dots) N/PMMoV WW data.

Figure 7 illustrates *R*_*e*_, which was computed assuming the median of the predicted PR multiplied by the average number of tests performed per day for Davis and UC Davis in the study period, *T* = 1, 198 and *T* = 2, 381 respectively. We also compare with the *R*_*e*_ computed with the observed cases. *R*_*e*_ trends determined from WW are similar in magnitude and depict similar trends for *R*_*e*_ calculated using observed cases during the periods analyzed.

**Figure 7:**
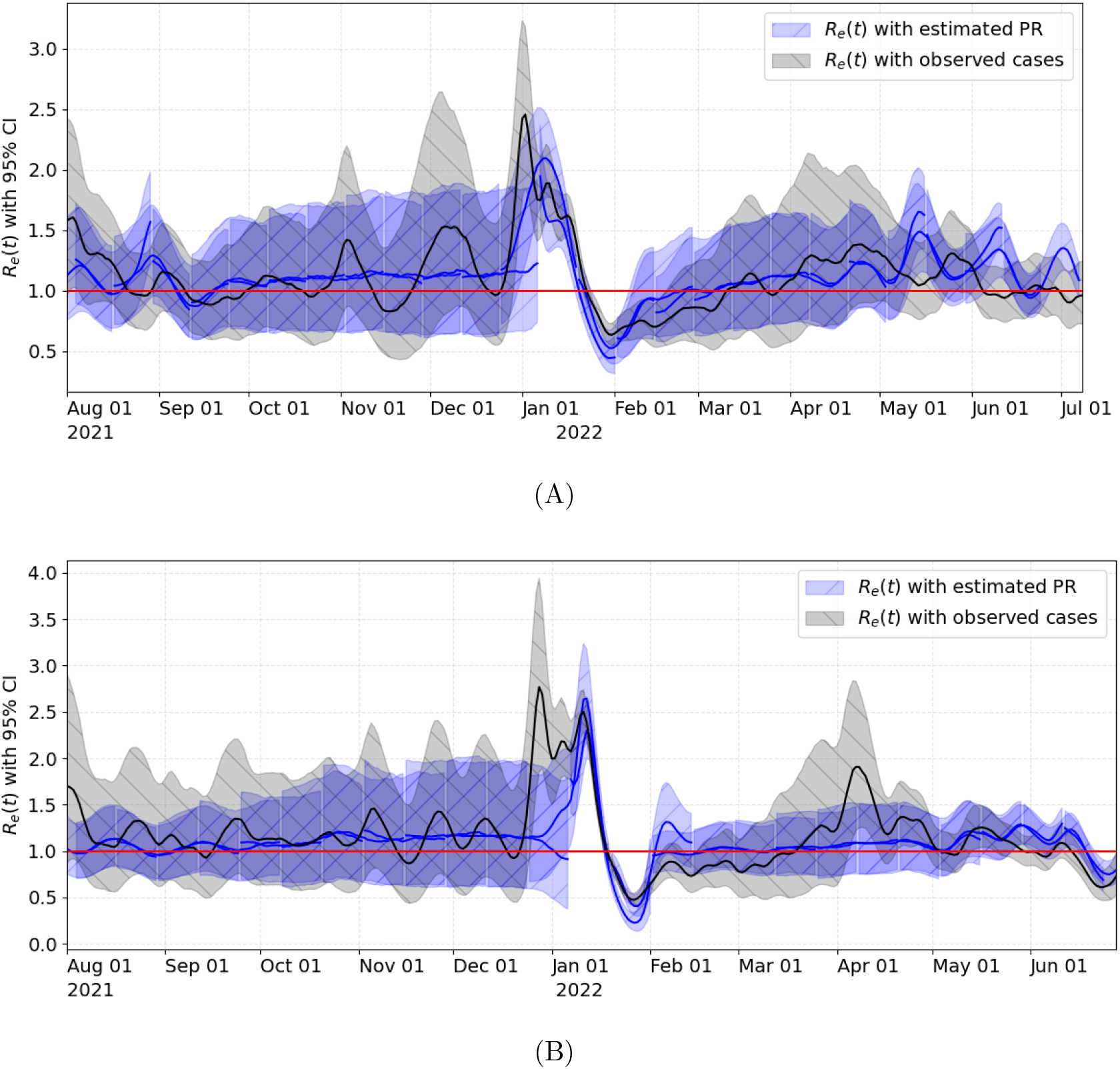
Effective *R*_*e*_ of (A) the City of Davis and (B) UC Davis computed with observed cases and the median of the predicted PR multiplied by the average number of tests performed for Davis in the study period. Solid lines and shaded regions illustrate the median and 95% prediction intervals, respectively.

## 4 Discussion

Substantial changes in test availability and test seeking behavior may confound estimates of case counts in a community (Daza-Torres et al., 2023). Positivity rate (PR) may provide a more accurate reflection of the state of the epidemic. PR is an important metric because it indicates how widespread an outbreak is within a particular area where testing is being conducted and whether current levels of testing are sufficient to accurately capture levels of disease transmission. Increases in PR can indicate that it may be a good time to incorporate restrictions to slow the spread of disease (Dowdy and D’Souza, 2020).

This study proposes a sequential Bayesian framework to model the COVID-19 PR via wastewater data for near real-time monitoring of the COVID-19 pandemic. The PR is modeled as a reparametrized Beta regression, and the parameters are estimated using a Bayesian approach. The changing dynamics of the virus and the data availability impose a challenge in developing helpful mathematical modeling for the surveillance and monitoring of COVID-19. Here, we propose an adaptive modeling framework as an alternative to overcome some of the limitations of traditional models. The adaptive capacity of the model is well suited to capture the variability in virus trends over time by leveraging knowledge gained.

We then use the model developed to offer a retrospective estimate of WW thresholds (for the settled solids analytical method performed) that corresponded to PR thresholds recommended by the CDC. The WW thresholds determined for UC Davis appeared more stable through time and through waves of different COVID-19 variants than WW thresholds estimated for the City of Davis. This can be explained by the fact that the UC Davis PR was nearly always less than 5%, while the City of Davis PR exceeded 5% periodically over the study period. Confidence in disease dynamics breaks down as PR rises above 5% (WHO, 2020). The WW thresholds estimated for the City of Davis were similar to those for UC Davis when PR remained low but increased dramatically at the end of the study period when clinical testing rates declined in Davis, usage of at-home test kits increased, and PR surpassed 5%. Mandatory asymptomatic testing continued for UC Davis through the end of the study period but has since been eliminated. In the absence of strong clinical testing programs, the relatively more stable WW thresholds determined over this study period may serve as a future reference to assess relative COVID-19 infection dynamics for these sewersheds. We caution against the direct translation of the estimated WW thresholds to other sewersheds and analytical methods for WW. Further research is needed to investigate the application of this framework to other sewersheds, for alternative WW analytical methods, and other respiratory and enteric pathogens present in WW.

One of the limitations of this framework is that it requires access to both WW and test data for continued adaptation, which implies continuous community monitoring through testing. The capacity of current testing programs has decreased significantly with recent transitions to a new normal and the implementation of prevention and surveillance mechanisms such as vaccines, at-home tests, and WW surveillance. New limited testing may lead to passive case-finding (i.e., only those most likely to be infected are tested). The PR may thus overestimate the current burden of the disease under these conditions. It is important to highlight that reduction in information poses new challenges and limitations in COVID-19 monitoring. With reductions in testing, public health authorities must decide between an indicator that overestimates the burden of the disease (PR) or a projection that underestimates the burden of the disease (case counts). We propose to use the PR in combination with WW measurements to reduce bias in assessments of disease dynamics.

In our study, the relationship between wastewater concentrations and PR changed when PR increased. This was likely due to changes in test-seeking behavior and test availability. Viral shedding may also change through time due to changes in vaccination status, acquired immunity, and changes in transmission patterns for different viral variants -although there is limited evidence available from fecal shedding studies. WW thresholds were estimated herein based on PR for a community where testing rates were extraordinarily high, given the population size. While it is not feasible at this time to establish *a priori* public health thresholds based on WW concentrations to estimate the burden of the disease through time, a record of historical values and thresholds provides meaningful context to guide public health authorities as new waves of infection arise.

## Data Availability

All data produced in the present study are available upon reasonable request to the authors.

https://github.com/mdazatorres/Bayesian_sequential_approach_PR_WW

## Availability Statement

All code and underlying data are publicly available through the GitHub repository (Montesinos-López and Daza-Torres, 2023). Analyses were carried out using Python version 3.9.

## Human Participant Protection

This study was determined to be exempt from institutional review board review by the UC Davis Office of Research.

## Credit authorship contribution statement

**J.C.M.L. and M.L.D.T**: Conceptualization, Methodology, Software, Formal analysis, Writing - Original draft preparation, Writing - Reviewing & Editing. **Y.E.G. and C.H**.: Literature review, Writing - Reviewing & Editing. **C.W.B**. Laboratory sample processing, Writing - methods. **H.N.B**.: Project conception, funding, research oversight, collaborator coordination, Writing - methods, Writing - Reviewing & Editing. **M.N**.: Methodology, research oversight/supervision, Writing - Reviewing & Editing.

## Declaration of competing interest

The authors declare that they have no known competing financial interests or personal relationships that could have appeared to influence the work reported in this paper.

## Acknowledgments

This research was supported by the National Center for Advancing Translational Sciences, National Institutes of Health, through grant number UL1 TR001860. The content is solely the responsibility of the authors and does not necessarily represent the official views of the NIH. This research was also supported by Healthy Davis Together (HDT) programs at the University of California, Davis, and the Centers for Disease Control and Prevention Foundation. Additionally, we thank Colleen C. Naughton, Alexandria B. Boehm, and Marlene K. Wolfe for their valuable comments. All authors reviewed and approved the final manuscript. We declare no conflicts of interest.

## Footnotes

1 The definition says that for given *p*, we are looking for some *y* (PR), such that *F* (*y*| *c*) *≥ p*. However, since multiple values of *y* could meet this condition, we take the smallest *y* of those.

## Notes

### Competing Interest Statement

The authors have declared no competing interest.

### Author Declarations

This study was determined to be exempt from institutional review board review by the University of California, Davis IRB Administration. IRB ID: 45 CFR 46.102.

### Summary of Updates

Corrected title: Missing an "r" in "wastewater. "wastewate" -> "wastewater".

